# The Breast Cancer Genetic Testing Experience: Probing the Potential Utility of an Online Decision Aid in Risk Perception and Decision Making

**DOI:** 10.1101/2024.09.13.24313647

**Authors:** Anna Vaynrub, Brian Salazar, Yilin Eileen Feng, Harry West, Alissa Michel, Subiksha Umakanth, Katherine D. Crew, Rita Kukafka

## Abstract

**Background:** Despite the role of pathogenic variants (PVs) in cancer predisposition genes conferring significantly increased risk of breast cancer (BC), uptake of genetic testing (GT) remains low, especially among ethnic minorities. Our prior study identified that a patient decision aid, *RealRisks,* improved patient-reported outcomes relative to standard educational materials. This study examined patients’ GT experience and its influence on subsequent actions. We also sought to identify areas for improvement in *RealRisks* that would expand its focus from improved GT decision-making to understanding results.

**Methods:** Women enrolled in the parent randomized controlled trial were recruited and interviewed. Demographic data was collected from surveys in the parent study. Interviews were conducted, transcribed, and coded to identify recurring themes. Descriptive statistics were generated to compare the interviewed subgroup to the original study cohort of 187 women.

**Results:** Of the 22 women interviewed, 11 (50%) had positive GT results, 2 (9.1%) with a *BRCA1/2* PV, and 9 (40.9%) with variants of uncertain significance (VUS). Median age was 40.5 years and 15 (71.4%) identified as non-Hispanic. Twenty (90.9%) reported a family history of BC, and 2 (9.1%) reported a family history of *BRCA1/2* PV. The emerging themes included a preference for structured communication of GT results and the need for more actionable knowledge to mitigate BC risk, especially among patients with VUS or negative results. Few patients reported lifestyle changes following the return of their results, although they did understand that their behaviors can impact their BC risk.

**Conclusions:** Patients preferred a structured explanation of their GT results to facilitate a more personal testing experience. While most did not change lifestyle behaviors in response to their GT results, there was a consistent call for further guidance following the initial discussion of GT results. Empowering patients, especially those with negative or VUS results, with the knowledge and context to internalize the implications of their results and form accurate risk perception represents a powerful opportunity to mediate subsequent risk management strategies. Informed by this study, future work will expand *RealRisks* to foster an accurate perception of GT results and include decision support to navigate concrete next steps.

## BACKGROUND

One of the primary bottlenecks to efficacious and inclusive breast cancer (BC) prevention is the identification of high-risk patients. Specifically, identifying women with pathogenic variants (PVs) in *BRCA1* and *BRCA2* (*BRCA1/2*) can inform risk management and prevention strategies to reduce the risk of developing breast and ovarian cancer^1, 2^.Women with hereditary breast and ovarian cancer syndrome (HBOC) attributable to *BRCA1/2* PVs have a lifetime BC risk of 40% to 60% and a lifetime ovarian cancer risk of 20% to 40%^3–5^. Additionally, up to 10% of breast cancers and 15% to 20% of ovarian cancers are attributed to PVs in HBOC predisposition genes [6]. Risk management strategies, including enhanced BC screening with mammography and breast MRI, chemoprevention, and risk-reducing surgeries such as prophylactic mastectomy or bilateral salpingo-oophorectomy (BSO) can significantly decrease a *BRCA1/2* carrier’s cancer risk (up to 90% with risk-reducing surgeries) once the patient is identified ^6–11^.

The United States Preventive Services Task Force (USPSTF) recommends that primary care providers (PCPs) screen asymptomatic women for increased risk of carrying *BRCA1/2* PVs^12, 13^. Pertinent risk factors include early onset of breast or ovarian cancer, multiple cases of breast or ovarian cancer in the family, bilateral breast cancer, male breast cancer, Ashkenazi Jewish descent, or a previously identified *BRCA1/2* PV in the family^13^. Although the indications and availability of genetic testing (GT) continue to expand, many women at an elevated risk of carrying *BRCA1/2* PVs are never identified^14–17^. Racial/ethnic minorities, along with patients of lower education and income levels, are less likely to be referred for GT, further perpetuating disparities in clinical outcomes^14, 18–20^.

Given the expanding criteria for GT, the need for genetic risk assessment continues to increase. One study found that less than 20% of patients had their genetic test ordered by a genetic counselor, and only approximately 50% of patients who underwent GT then discussed their results with a genetic counselor^18^. Given the limited accessibility of genetic counselors to even those patients with clinical indications for GT, decision support tools may offer an alternative for average-risk patients by providing similar information to that communicated in counseling sessions while preserving counseling resources for higher-risk patients^18^. Establishing the effectiveness of alternative strategies for both pre-test counseling and return of results will be crucial to addressing the increased need for GT services as more patients are identified for BC genetic risk assessment.

Various studies have shown that risk perception, potentially more so than the GT result itself, may significantly influence patient’s medical decision-making and associated clinical outcomes^21, 22^. However, it is crucial to recognize that the delivery of information does not equate to neither knowledge retention nor accurate risk perception^21^. These trends suggest that the way forward is to meet the patients at this level of discrepancy and introduce tools that help facilitate more accurate risk perception and provide support at every step of the diagnostic process.

To this end, Kukafka *et al.* developed and evaluated the web-based *RealRisks* decision aid (DA) for women to screen for GT eligibility and a complementary decision support tool called Breast Cancer Risk Navigation Tool (*BNAV*) for their primary care providers (PCPs)^23^. In a cluster randomized controlled trial (RCT) of standard educational materials alone vs. in combination with *RealRisks* and *BNAV* among 187 women and 67 clinicians, respectively, there was a significant decrease in BC worry and perceived lifetime BC risk in the intervention compared to the control arm ^23^. However, there was no significant increase in the primary endpoint of genetic counseling uptake at 6 months in the intervention vs control arm (19.8% vs. 11.6%, p=0.14) ^23^. These similar rates of GC uptake may reflect the very decrease in BC worry conferred by *RealRisks* engagement.

Research to understand the experiences of women who underwent HBOC GT following exposure to DAs such as *RealRisks* is limited. Therefore, we conducted semi-structured interviews to identify how women enrolled in the RCT who opted for GT understood, interpreted, and acted upon their GT results. The aims of this study include: 1) examining patients’ GT experience and its influence on subsequent actions pertaining to BC prevention and follow-up; and 2) identifying areas for improvement in *RealRisks* that would expand its focus from improved GT decision-making (*e.g.,* the decision to test) to understanding and interpreting results from HBOC GT (*e.g.,* return of results).

## METHODOLOGY

### Recruitment

Individuals were recruited from the cohort of the parent study^23^. Eligible patients were age 21-75 years, without a personal history of breast or ovarian cancer, no history of genetic counseling or testing for HBOC, eligible for *BRCA1/2* GT based on a validated family history screener^18^, and ability to provide informed consent in English or Spanish ^23^. Of 187 evaluable patients (101 in the intervention group and 86 in the control group), a total of 58 patients had received GT confirmed by the electronic health record (EHR) at 24 months following study enrollment and were eligible for this qualitative nested study. Contact information from enrollment in the prior study was utilized for recruitment. Outreach was conducted by email and phone. Study procedures were approved by the Columbia University Irving Medical Center (CUIMC) Institutional Review Board. Patient eligibility and recruitment are detailed in **Figure 1**.

**Figure 1.**
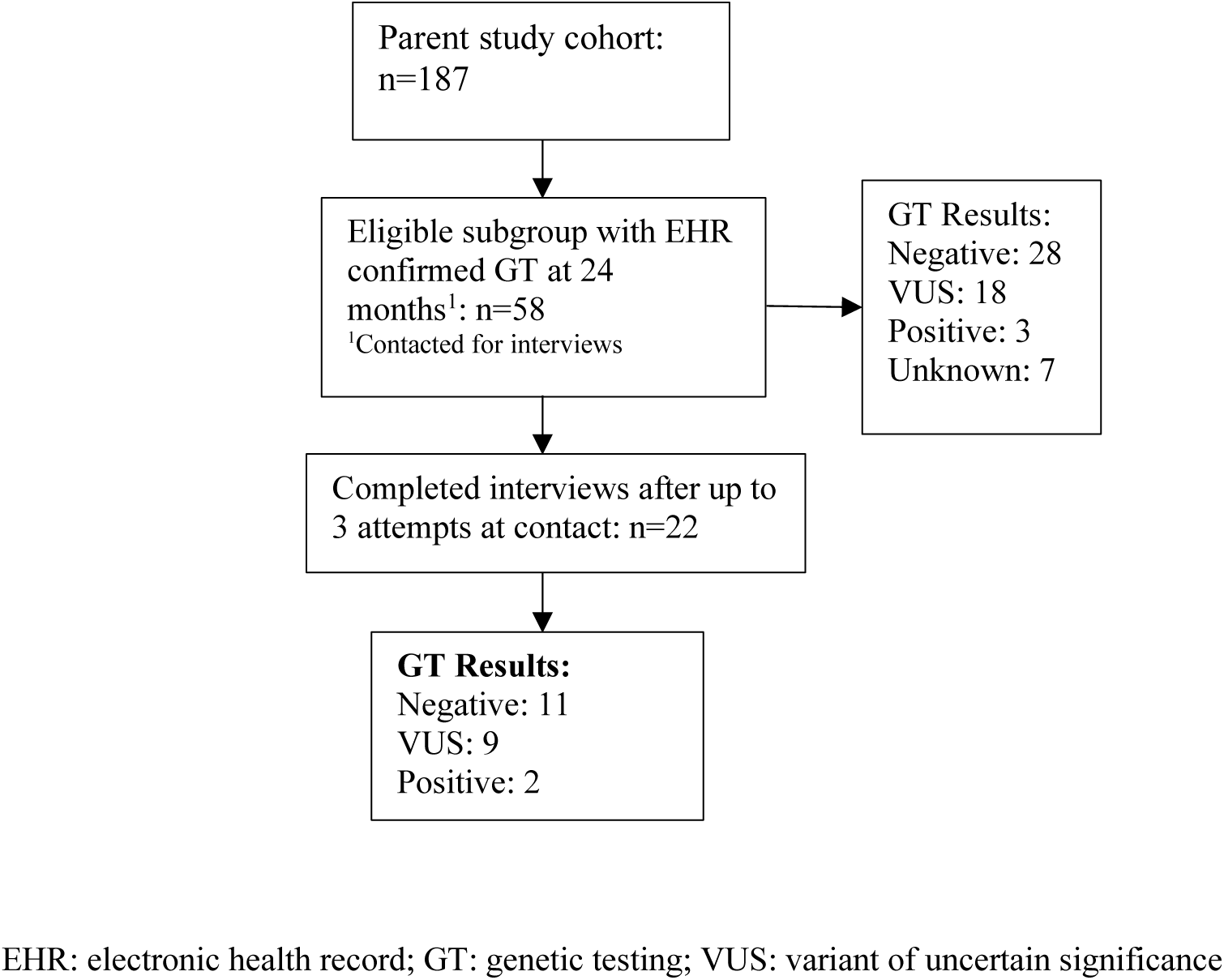
Cohort Selection Flow Diagram

### Data Collection

Two versions of the interview guide were developed corresponding to the participant’s original assignment to the control or intervention arm in the parent study (**Appendix A).** Participants were further grouped based on their GT results: negative, positive, or variants of uncertain significance (VUS). Patients were categorized as having “unknown” results if there was no available documentation of their results and they were unable to corroborate their results themselves. Interviews were conducted in Spanish and English by a bilingual interviewer. The interviews took place over Zoom video conference and were audio recorded and transcribed. Demographic and BC risk factor data for each participant were collected from surveys administered during the parent study.

### Codebook and Qualitative Data Analysis

The research team developed a codebook to identify themes that could provide insight into the experiences of women who underwent HBOC GT. Each code was assigned a definition with instructions regarding code application. The coding team was comprised of two experienced coders. Scott’s Pi was tabulated as a measure of intercoder reliability at 0.624, indicating substantial agreement ^24^. The qualitative data analysis platform ATLAS.ti. was used to code transcripts by the research team.

### Quantitative Statistical Analysis

To compare the interviewed subgroup to the total eligible cohort, comparison of demographic characteristics was conducted using R Studio (Version 1.4.1717). Normality was determined using the Shapiro-Wilks normality test. Data for continuous variables are presented as medians (with interquartile range) and counts and percentages for categorical variables. The Kruskal-Wallis test and ANOVA were used to compare variables as appropriate. Categorical variables were compared using the Pearson chi-squared test. A p-value less than 0.05 was deemed significant for all statistical analyses.

## RESULTS

Of the 58 patients recruited from the parent trial who were eligible for this study, 22 (38%) were interviewed. As shown in **Table 1**, a comparison of the interviewed patients to the total cohort revealed minimal inter-group differences except for family history of *BRCA1/2* pathogenic variants (PVs). Of the 58 eligible patients, 29 (50%) had negative GT results, 19 (33%) had a variant of unknown significance (VUS), 3 (5%) had a PV, and 7 (12%) had unknown results (**Table 1**). Of the 22 participants who completed the interview, 6 (27%) had VUS results, 2 (9%) had a PV, and 11 (50%) had negative results. Half of those interviewed (n=11; 50.0%) identified as Jewish. Self-reported racial identities of the interviewed participants included White (n=16; 72.7%), Black/African American (n=5; 22.7%), and Asian (n=1; 4.5%). All participants had completed their high school education at a minimum.

**Table 1.**
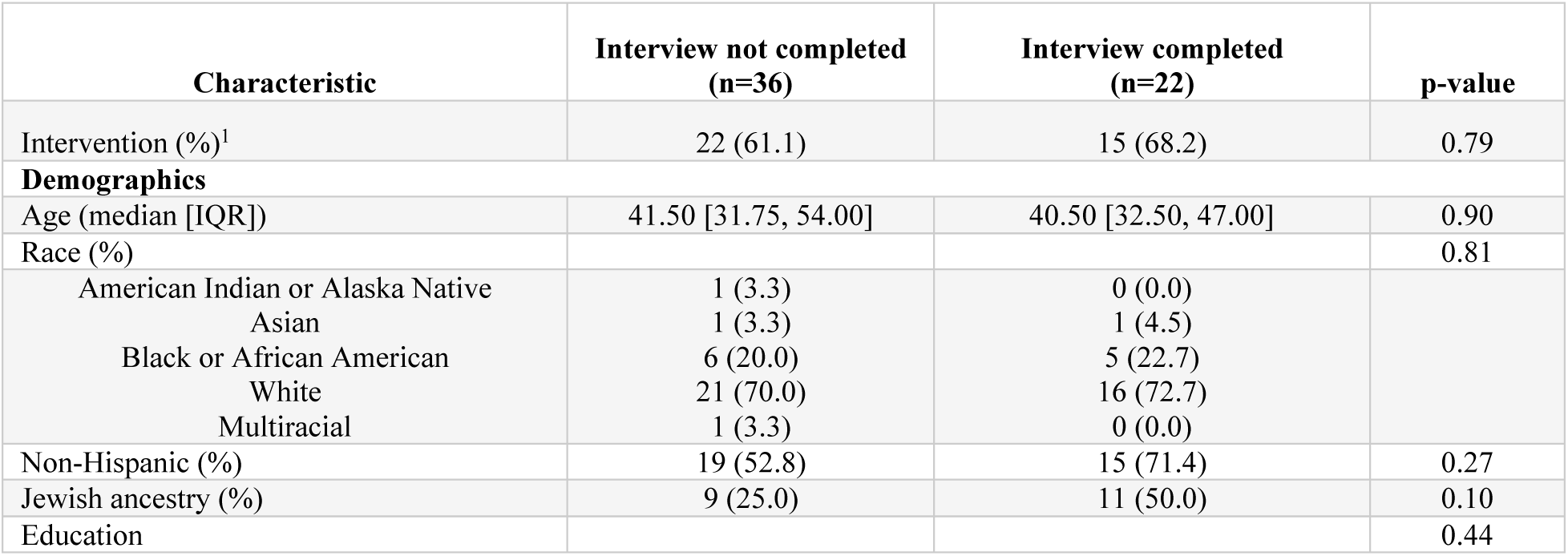
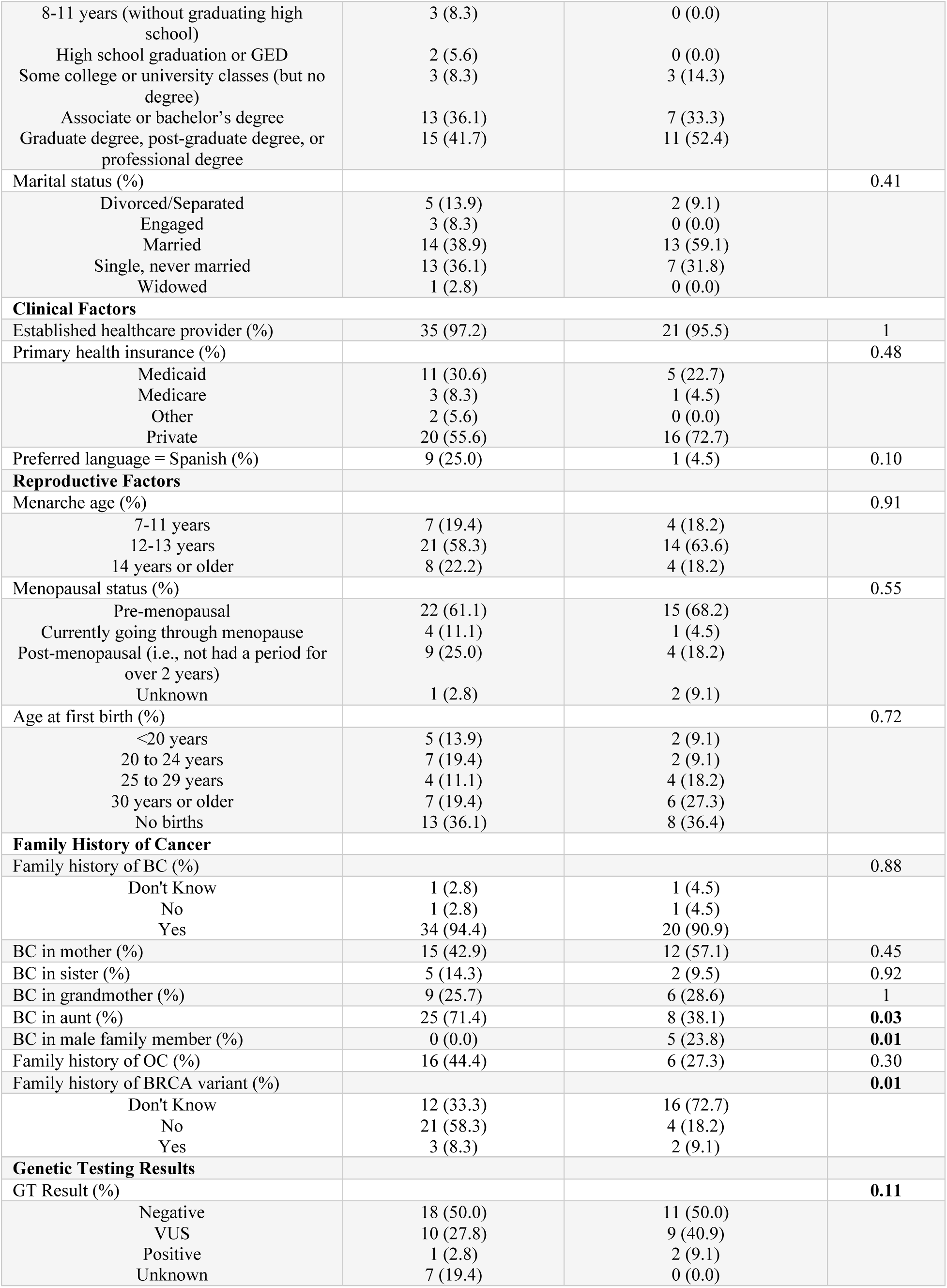
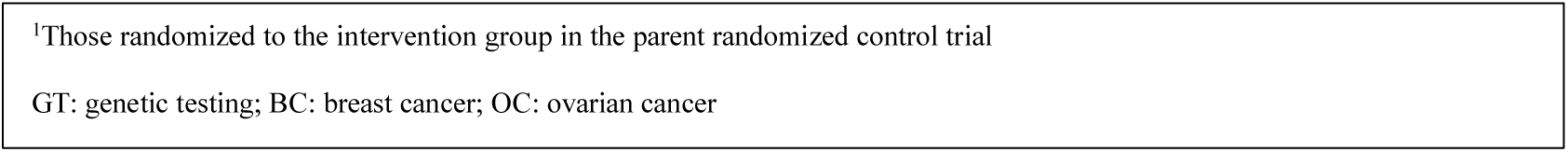
Baseline participant characteristics stratified by interview participation.

| Characteristic | Interview not completed<br>(n=36) | Interview completed<br>(n=22) | p-value |
| --- | --- | --- | --- |
| Intervention (%)<sup>1</sup> | 22 (61.1) | 15 (68.2) | 0.79 |
| **Demographics** |  |  |  |
| Age (median [IQR]) | 41.50 [31.75, 54.00] | 40.50 [32.50, 47.00] | 0.90 |
| Race (%) |  |  | 0.81 |
| American Indian or Alaska Native | 1 (3.3) | 0 (0.0) |  |
| Asian | 1 (3.3) | 1 (4.5) |  |
| Black or African American | 6 (20.0) | 5 (22.7) |  |
| White | 21 (70.0) | 16 (72.7) |  |
| Multiracial | 1 (3.3) | 0 (0.0) |  |
| Non-Hispanic (%) | 19 (52.8) | 15 (71.4) | 0.27 |
| Jewish ancestry (%) | 9 (25.0) | 11 (50.0) | 0.10 |
| Education |  |  | 0.44 |
| 8-11 years (without graduating high school) | 3 (8.3) | 0 (0.0) |  |
| High school graduation or GED | 2 (5.6) | 0 (0.0) |  |
| Some college or university classes (but no degree) | 3 (8.3) | 3 (14.3) |  |
| Associate or bachelor's degree | 13 (36.1) | 7 (33.3) |  |
| Graduate degree, post-graduate degree, or professional degree | 15 (41.7) | 11 (52.4) |  |
| Marital status (%) |  |  | 0.41 |
| Divorced/Separated | 5 (13.9) | 2 (9.1) |  |
| Engaged | 3 (8.3) | 0 (0.0) |  |
| Married | 14 (38.9) | 13 (59.1) |  |
| Single, never married | 13 (36.1) | 7 (31.8) |  |
| Widowed | 1 (2.8) | 0 (0.0) |  |
| **Clinical Factors** |  |  |  |
| Established healthcare provider (%) | 35 (97.2) | 21 (95.5) | 1 |
| Primary health insurance (%) |  |  | 0.48 |
| Medicaid | 11 (30.6) | 5 (22.7) |  |
| Medicare | 3 (8.3) | 1 (4.5) |  |
| Other | 2 (5.6) | 0 (0.0) |  |
| Private | 20 (55.6) | 16 (72.7) |  |
| Preferred language = Spanish (%) | 9 (25.0) | 1 (4.5) | 0.10 |
| **Reproductive Factors** |  |  |  |
| Menarche age (%) |  |  | 0.91 |
| 7-11 years | 7 (19.4) | 4 (18.2) |  |
| 12-13 years | 21 (58.3) | 14 (63.6) |  |
| 14 years or older | 8 (22.2) | 4 (18.2) |  |
| Menopausal status (%) |  |  | 0.55 |
| Pre-menopausal | 22 (61.1) | 15 (68.2) |  |
| Currently going through menopause | 4 (11.1) | 1 (4.5) |  |
| Post-menopausal (i.e., not had a period for over 2 years) | 9 (25.0) | 4 (18.2) |  |
| Unknown | 1 (2.8) | 2 (9.1) |  |
| Age at first birth (%) |  |  | 0.72 |
| <20 years | 5 (13.9) | 2 (9.1) |  |
| 20 to 24 years | 7 (19.4) | 2 (9.1) |  |
| 25 to 29 years | 4 (11.1) | 4 (18.2) |  |
| 30 years or older | 7 (19.4) | 6 (27.3) |  |
| No births | 13 (36.1) | 8 (36.4) |  |
| **Family History of Cancer** |  |  |  |
| Family history of BC (%) |  |  | 0.88 |
| Don't Know | 1 (2.8) | 1 (4.5) |  |
| No | 1 (2.8) | 1 (4.5) |  |
| Yes | 34 (94.4) | 20 (90.9) |  |
| BC in mother (%) | 15 (42.9) | 12 (57.1) | 0.45 |
| BC in sister (%) | 5 (14.3) | 2 (9.5) | 0.92 |
| BC in grandmother (%) | 9 (25.7) | 6 (28.6) | 1 |
| BC in aunt (%) | 25 (71.4) | 8 (38.1) | **0.03** |
| BC in male family member (%) | 0 (0.0) | 5 (23.8) | **0.01** |
| Family history of OC (%) | 16 (44.4) | 6 (27.3) | 0.30 |
| Family history of BRCA variant (%) |  |  | **0.01** |
| Don't Know | 12 (33.3) | 16 (72.7) |  |
| No | 21 (58.3) | 4 (18.2) |  |
| Yes | 3 (8.3) | 2 (9.1) |  |
| **Genetic Testing Results** |  |  |  |
| GT Result (%) |  |  | **0.11** |
| Negative | 18 (50.0) | 11 (50.0) |  |
| VUS | 10 (27.8) | 9 (40.9) |  |
| Positive | 1 (2.8) | 2 (9.1) |  |
| Unknown | 7 (19.4) | 0 (0.0) |  |

<sup>1</sup>Those randomized to the intervention group in the parent randomized control trial
GT: genetic testing; BC: breast cancer; OC: ovarian cancer

### Qualitative Results

Interviews were initially analyzed using the following nine codes: 1) behavioral changes based on GT results, 2) communication of GT results, 3) experience receiving results, 4) initial reaction to GT results, 5) understanding/lack of understanding of GT results, 6) method of receiving GT results, 7) *RealRisk* references, 8) recommendations regarding GT, and 9) understanding of risk factors. **Table 2** documents the frequency with which each code was applied across the interview transcripts. Five themes emerged from the transcripts: 1) preferences regarding communication of GT results; 2) lifestyle changes influenced by GT; 3) understanding and emotional reception of GT results; 4) utility and role of *RealRisks* in deciding to pursue GT; and 5) recommendations on how to improve the GT process.

**Table 2:** Frequency of code use across interview transcripts.

| Code | Frequency |
| --- | --- |
| Behavioral changes based on GT result | 109 |
| Communication of GT results | 64 |
| Experience receiving GT results | 137 |
| Initial reaction to GT results | 31 |
| Understanding/ lack of understanding of GT results | 75 |
| Method of receiving GT results | 39 |
| *RealRisk* references | 59 |
| Recommendations regarding GT testing | 37 |
| Understanding of risk factors | 84 |

GT: genetic testing

### Theme 1: Preferences regarding communication of GT results

Regardless of the classification of the GT result, patients frequently expressed appreciation for the role of genetic counselors and particularly the opportunity to discuss their GT results in a face-to-face encounter. Among patients who provided a direct answer to “who was most helpful” (n=9), genetic counselors and medical professionals were considered to be the “most helpful” in explaining their GT results (33.3% and 55.6%, respectively). In contrast, receiving results, even from a genetic counselor, over the phone was viewed as a stressful event. Comments associated with understanding or having few questions after receiving results were frequently associated with having worked with a genetic counselor in person in the setting of a structured visit. As one patient summarized:

> *“Well, I did follow up, you know, when I met with the counselor and they also suggested seeing somebody at the hospital, which I did. They were very informative.”*

Patients who received PV results specifically cited that delivery of results over the phone was not favorable (n=2). One of the participants expressed the following:

> *“It was pretty bad. I think hearing that by phone was definitely not what I was expecting. I was in the middle of a workday, and I got this random call, and this person started telling me all these things. That was a little disappointing, I think for me.”*

Another patient who had received notification of a PV over the phone similarly noted:

> *“I think for me it was 2 things. First, I think if I had an appointment where this would be told to me in person and I had like time to process the information and go over things, it would have been much easier.”*

As one patient with a negative result alluded to, the negative emotions surrounding receiving positive results may be overwhelming, suggesting that an in-person approach may be favorable:

> *“I do understand that some people can get so clouded by the fact they possibly could have cancer, they may not even digest what they’re telling you but my recommendation for sure is definitely keep doing the in-person, sitting down with the person and maybe holding their hand through the process of really understanding what that genetic test can be.”*

Whereas patients with PVs noted dissatisfaction with the brevity and lack of support in receiving their results, especially by phone, patients with VUS and negative results more often expressed appreciation of the depth and time taken to fully explain their results to them. Moreover, those with VUS and negative results reported an overall more positive experience in receiving their GT results. One patient who had received the result of VUS explained:

> *“I think again having a counselor explain what they did, what the results mean, where does that put you in a risk bracket, it’s highly helpful for me as a patient to know that okay, I know that maybe I don’t need to take additional drastic changes or actions, but I should continue to do what I need to do, to go to the doctor to check every year, mammograms, and I know that hopefully the genetics company is going to update me if new information or new technology changes the view of the results.”*

Overall, patients with PVs consistently reported a perceived benefit with face-to-face genetic counselor sessions, and patients with VUS or negative results were more often satisfied with the communication they received relative to patients who received PV results.

### Theme 2: Lifestyle changes influenced by GT

While most (n=14, 65%) patients reported no changes in daily behaviors or medical management in response to their results, those that did most commonly described changes in diet, exercise, screening, and smoking. These lifestyle changes were reported by patients who received positive and negative results. Commonly identified risk factors among participants, regardless of GT result, were alcohol use, smoking, and family history. As one patient with a PV result described:

> *“I feel like there are so many epigenetic factors, so that comes into daily life and it’s really made me more conscious about everything from eating well to sleeping well to exercising to making sure I follow through on some of the screening recommendations, so yeah I think it has had an impact on my daily life”*

Both patients with PVs reported that receiving their results influenced family planning, with one patient stating:

> *“…we did go through IVF to select for non-BRCA genes in the embryos so yeah it had an impact on my family”*.

While most patients did not report lifestyle changes following the return of their GT results, most did understand that their behaviors, such as sleeping, exercise, and diet, can impact their BC risk.

### Theme 3: Understanding and emotional reception of GT results

In discussing the implications of their GT results, women expressed an understanding that negative GT or the lack of a PV does not guarantee a cancer-free lifetime (n=15, 75%). Patients who had received negative or VUS results commonly reported more positive emotions receiving their results compared to those with PVs. Nearly half of the VUS patients (44.4%) explicitly reported feeling relief on receiving their results. All patients with a negative result described their feelings as related to relief and joy (n=11 out of 11, 100%):

> “*It was great. I was ecstatic. She was very happy. I was happy.”;*

> *“When you have a family history of cancer, it’s scary to think about and you never know what’s in your future, so just the fact that I’m not predisposed to it was a little bit of a relief.”*

Family experience with either cancer or GT was noted as influencing the patient’s experience of receiving and processing GT results by five patients (22.7%). There was no explicit pattern regarding the nature of this influence based on the GT result. Some patients described family experience as positive, pointing towards the ability to tap into a sense of familiarity, support, and perception of information access (“…*helpful in providing some information”*). Instances in which family experience contributed negatively centered around increasing anxiety due to either apprehension of risk or healthcare experience. For one patient who received a positive result, prior family experience with cancer offered some reassurance:

> *“I think like the reason why I was not scared was technically because I knew I was young, my mom had cancer when she was young and they found it right away and she was treated. So, I felt empowered that I knew I had this predisposition but I was aware of it and I could act upon.”*

In contrast, another patient with a negative result explained:

> *“I guess I was more suspicious that I had to come in, I guess. I had an experience with genetic testing for another family member and it was kind of like if it was negative, you just found out and if it was positive, you had to come in so I think it provoked more anxiety than maybe just getting the test results however and following up.”*

Regardless of their results, patients expressed strong emotions, which varied from relief to anxiety, and it was evident that personal experience, especially with family members, can impact emotional responses.

### Theme 4: Utility and role of *RealRisks* in deciding to pursue GT

Just over one-third of patients enrolled in this study reported that they had formed intentions to undergo GT prior to engaging with *RealRisks*. Specifically, eight participants of those interviewed (36.4%) stated that *RealRisks* did not have any impact on their decision to pursue GT as they had already made the decision to do so prior to accessing *RealRisks*. Comments such as: “*It didn’t change my decision because I was going into it with the intention of getting the actual test*” were common. This sentiment was consistent across GT results.

Although the preformed decision to pursue GT is unsurprising given this self-selected patient population, *RealRisks* may assist in contemplating the GT decision and in understanding BC risk factors beyond PVs.

### Theme 5: Recommendations on how to improve the GT process

Across GT results, patients frequently cited the need for more guidance regarding next steps after receiving their results. Specifically, over one-third of patients felt more information was needed regarding modifiable lifestyle factors and action items that can be implemented to reduce risk. These patients described a lack of GT awareness, understanding of risk factors, and education regarding the next steps. One participant commented:

> *“I feel that I mean I probably should be able to describe in detail what the known risk factors are, but I don’t know that I could.”*

Another patient described:

> *“A lot of questions opened up about what were the implications of that, like I knew the overall implications, but I had a lot of questions about the details about what would be the changes to my sort of day-to-day life and when would I have to start preventative screening and those things.”*

One specific comment expressed that this need is especially underscored among the Latino/Hispanic community:

> *“..it would help if the genetic counselor, the website [RealRisks] and the doctors empower the patient with more information in terms of what they can do for their lifestyle, even if it’s not proven, especially in the Latino/Hispanic community, they are looking for solutions on what they can do.”*

On a similar note, participants (n=2 out of 22, 9.1%) recommended that there be an increase in awareness of testing availability among patients, in so doing expanding the accessibility of GT. Specifically, one Latinx patient stated that:

> *“I think that a lot of people, even my own peers and my own community, don’t have an understanding of the importance of getting genetic testing, the access to it, the fact that in addition to evolution in the medical community, insurance has improved and kind of financial accessibility to it. Although I don’t know that I necessarily would benefit from it, and it almost feels like more publicity, but more global and communal awareness around genetic testing. I do believe I’m part of a community and my larger community would certainly benefit from that.”*

In summary, explanations of actionable follow-up and risk factors are among the most common points of feedback.

## DISCUSSION

This qualitative interview study aimed to explore the salient factors that influenced the genetic testing (GT) experiences of women who participated in a decision support intervention to increase appropriate *BRCA1/2* GT in the primary care setting. *RealRisks,* a patient-facing web-based decision aid (DA), was designed to improve a patient’s knowledge of breast cancer (BC) and personalized risk, as well as to support GT decision-making. As we plan to expand *RealRisks*, another goal of this interview study was to understand how *RealRisks* can inform every step of the GT experience, from risk assessment, education, decision support, return of results, and follow-up care and lifestyle modification.

Following the initial risk assessment to determine eligibility for GT, patients participating in the parent randomized controlled trial (RCT) viewed *RealRisks* as a tool for making an informed decision in pursuing or declining GT. In the RCT, *RealRisks* significantly reduced BC worry and helped inform BC risk perceptions compared to the control groups that received standard educational materials. This suggests that even when the intention to receive GT may be formed independently of *RealRisks*, as was the case with most participants interviewed in this nested study, the *RealRisks* DA may be useful in improving measures of decision quality. Additionally, translating intention to actual behavior (e.g., to pursue GT when indicated) requires favorable attitudes and perceived behavioral control, which are modifiable factors that could be targeted within *RealRisks* to help at-risk individuals recognize their risk status, pursue appropriate GT, and engage in appropriate risk-mitigating actions^25^. It is also worth noting that in the parent study, the extent or “dose,” in terms of time spent using *RealRisks*, in contrast to exposure in itself, did correlate with GT uptake. This trend may also reflect the possibility that patients who were alerted to their risk through an external source (e.g., mammogram result, family history, etc.) were more motivated to thoroughly engage with *RealRisks* and pursue GT. Given that prior studies have established risk perception as a primary mediator of medical decision-making, these findings indicate that there exists a role for *RealRisks* in influencing GT decisions and motivating risk-mitigating actions; however, an adequate “dose” or extent of utilization is necessary.

After making the decision to pursue or decline GT, both understanding of testing results and the emotional context in which results are received are critical influencers of subsequent action. Currently, *RealRisks* does not include a return of results module. However, preparing the patient to receive and further act on their results, regardless of the result itself, is arguably as crucial as providing the result. Unlike the GT result itself—which has been found to have varying significance independently—the patient’s risk perception has been consistently found to be directly associated with the patient’s medical decision-making (including the decision to undergo prophylactic surgery)^21, 26^. Thus, empowering the patient with the knowledge and context with which to internalize the implications of their results and form accurate risk perception represents a powerful opportunity to mediate subsequent health behaviors.

Among patients who received negative test results, most understood that this result does not guarantee that they will never develop cancer. In this regard, patients with negative test results demonstrated an adequate understanding of lifetime risk and the role of non-genetic factors in contributing to risk. Nevertheless, the most frequently reported emotions surrounding variant of uncertain significance (VUS) and negative test results were joy and relief. Prior studies identified BC history and prior experience with cancer at large in the healthcare setting were also associated with emotional reception of GT results^27^. However, no significant trends in these factors influencing reaction to results were identified in this cohort.

Among patients dissatisfied with the communication of their GT results, specific complaints included feeling poorly supported, abrupt delivery of information, and requiring more guidance for subsequent steps in medical management. While positive experiences tended to correlate with in-person encounters and negative experiences with phone encounters, these patterns may also be a testament to the nature of these discussions. Those who were dissatisfied with result communication over the phone primarily touched on the abruptness and brevity of the encounter. In contrast, those who had positive experiences specifically mentioned the time taken to explain the patients’ risk based on their personal information. This finding may support the need for additional services, especially for patients with negative or VUS results given the frequency with which this group expressed the need for further guidance after receiving their results. Indeed, an RCT led by Molina *et al*. found that low-intensity navigation services increased the odds of subsequent screenings among women with negative or VUS results^28^. Interestingly, a recent survey examining communication preferences among various demographics found that non-Hispanic Black and older women preferred less detailed results communication^29^. While several patients in this study specifically emphasized their preference for in-person communication, the aforementioned study also found that those who preferred in-person communication were of higher risk while those of average risk preferred written letters^29^. Additionally, the limited number of patients in this study who voiced discontent with over-the-phone results delivery prevents generalizability of this trend to the larger target patient demographic.

Preferences for delivery of results are mixed and personal; nevertheless, patients frequently expressed appreciation of an in-person conversation with a genetic counselor in understanding their results. However, certain barriers, including a shortage of GT counselors and a shift to virtual appointments after the COVID-19 pandemic, have highlighted the need for alternative resources and methods. Indeed, the U.S. Genetic Counselor Workforce Working Group determined that a shortage of genetic counselors may persist through 2030^30^. This study, therefore, will serve to inform future work to address the persistent need for alternative personalized, accessible genetic service resources^31^ ^32^ ^33–35^.

Another particularly relevant theme that emerged was the need for more guidance regarding the next steps after receiving GT results, with several mentions of an experience in which a patient was presented with their GT result but felt unsure regarding next steps. In this study, this uncertainty was primarily identified in participants with negative results. Namely, a recurrent thread centered around patients voicing a need for both information and clarity regarding medical follow-up and daily risk reduction practices, especially in the Latinx/Hispanic community. While the relationship between race/ethnicity and BC knowledge has been contested across literature, there have been indications that race/ethnicity is, at the very least, a relevant factor in predicting patient activation, testing, and follow-up. Specifically, a 2015 single center survey study by Hong *et al.* found significant interaction effects between race and behavioral causal perceptions on cancer risk perception among African Americans compared to other races^36^. Additionally, a recent sequential mixed-methods study found that taken cumulatively, non-Hispanic Whites and individuals with greater health literacy at baseline had a more accurate understanding of their BC risk^37^. Moreover, few studies have focused on providing support for the management of patients who test negative for a PV but otherwise remain relatively high risk^38^.

While most patients in our study reported no change in behaviors in response to GT results, those who did cited diet, exercise, cancer screening, or substance use. Accordingly, when asked to describe known BC risk factors, patients also commonly referenced weight, dietary habits, smoking/drug use, and family history. These patterns suggest that patients are willing to apply their understanding of risk factors to actionable lifestyle changes. Interestingly, both patients with positive and negative results reported being motivated to implement lifestyle changes where applicable, indicating that the experience of GT itself rather than the results may serve as an impetus in behavior modification. These findings underscore the importance of providing the knowledge required to implement these changes, especially since numerous respondents self-reported a worrisome knowledge gap.

Patients with identified PVs tended to report a less thorough and supportive GT experience. This trend may indicate a paradoxical tendency to focus more on ensuring negative/VUS patients adequately understand the nuances of their results rather than the sensitivity of communicating positive results. This aligns with prior studies in which patients have a more accurate perception of positive results, perhaps due to their more apparent or ostensibly straightforward implications^39^.

Overall, the literature suggests that the problem of under-utilization of GT is complex, with barriers at multiple levels, and that there needs to be more widely implemented, long-term interventions to address this issue. While it may seem intuitive to focus on the breadth of genetic counseling/education services to increase GT uptake, various studies have shown that knowledge gain and retention are relatively limited after counseling^40, 41^. Thus, while there is a call for better communication of genetic risk via counseling, multiple studies have shown that it is the individual perception of risk, even given adequate risk communication, that more directly influences results^21, 26, 40^. Therefore, accounting for factors that impact risk perception in conjunction with information delivery may promote health equity in GT and improve clinical outcomes.

Regarding the applicability of *RealRisks*, the next step forward in bridging GT and appropriate downstream preventive services will be establishing a return of results module directly informed by the content of this interview study. This feature will provide the patient with critical context as to the medical implications of the participant’s result, along with specific, targeted action items. These items will include preventative measures, risk reduction strategies, and specific steps for follow-up where necessary.

We acknowledge several limitations to our study. Namely, the small size of the interviewed cohort limits the scope of experiences and perspectives solicited, thus limiting the generalizability of our findings. Additionally, all participants who were interviewed had decided to pursue GT prior to engaging with *RealRisks*, preventing us from obtaining an accurate understanding of the role of *RealRisks* in the initial decision to pursue GT. Moreover, the interviewed subgroup was enriched for patients with a family history of breast cancer (90.9%), potentially skewing the perspectives represented. Strengths of our study include quantitative data to better understand the relevant clinical and demographic context of our patient population. Additionally, the qualitative analysis was conducted by an interdisciplinary team of researchers who lent their respective expertise in medicine, public health, and biomedical informatics to thoroughly analyze and parse the interview data.

## Conclusion

Overall, we found that patients expressed preferences for a scheduled verbal explanation of their GT results to facilitate a more personal, supportive testing experience. Understanding of the implications of negative results was largely adequate. While most patients did not change lifestyle behaviors in response to their GT results, there was a consistent call for further guidance and navigation following the initial discussion of GT results. Thus, future efforts to enhance appropriate GT uptake and risk screening should focus on fostering an accurate perception of GT results and support in navigating concrete next steps.

## Data Availability

All data produced in the present study are available upon reasonable request to the authors.

## Declarations

### Ethics approval and consent to participate

Study materials and procedures were approved by the Columbia University Irving Medical Center (CUIMC) Institutional Review Board. Individual informed consent was granted by interviewed study participants through an online informed consent form.

### Consent for publication

Not applicable.

### Availability of data and materials

The datasets used and/or analyzed during the current study are available from the corresponding author on reasonable request.

### Competing interests

Not applicable.

### Funding

Funding was provided to RK by the American Cancer Society, grant number: RSG-17-103-01.

### Authors’ contributions

AV wrote the primary manuscript text, completed quantitative analysis, produced all tables and figures, and contributed to qualitative analysis of patient interviews. BS conducted patient interviews, coded interview transcripts, and contributed to qualitative analysis of patient interviews. YEF, HW, AM, and SU contributed to qualitative analysis and revision of the manuscript. KC and RK oversaw the completion of the study, guided development of methodology and results analysis, and provided revisions of the manuscript.

## Acknowledgements

Not applicable.

## Notes

### Competing Interest Statement

The authors have declared no competing interest.

### Clinical Trial

NCT03470402

### Author Declarations

IRB of Columbia University Medical Center gave ethical approval of this work.

