## Appendix A for "The Breast Cancer Genetic Testing Experience: Probing the Potential Utility of an Online Decision Aid in Risk Perception and Decision Making"

Interview Guide

*First, thank participant for taking part in the genetic counseling/test study and for*

*agreeing to participate in this interview.*

*Begin with:*

• Have you received a major diagnosis such as breast cancer since you

participated in this study? (Use EPIC data to see what month/year they got

genetic testing)

• Think back to when you received your genetic test result, what was your result

called? (If unsure, you can remind participants of the possible results, did you

receive a positive result, a negative result, or a VUS?)

• What does [insert result] mean to you?

• What does your [insert result] mean to your risk to develop breast cancer?

- How did you get your genetic test result?

• Did someone contact you to discuss your result with you?

• Did you have to contact your provider’s office (or genetic counselor’s)?

• If not, why not?

• What was that experience like for you?

Now, I’d like to talk to you about the conversation you had with [insert provider

type]. *(Ask this series of questions for all conversations with provider and/or genetic counselor).*

• What did you understand from the conversation you had?

• What did you not understand?

• Did you have any questions for the person who gave you your results?

• What, if anything, did you know about the possible test results you could receive

before actually receiving your result?

• How could we improve your knowledge of genetic test results?

• What was your initial reaction to your genetic test result?

• How did you feel when you heard your results?

• Relieved? Worried? Confused?

• What do you think influenced the way you reacted to your genetic test result (i.e., your family history, the genetic counseling you received)?

• Who has been the most helpful to you to understanding your genetic testing? (i.e., friend, family ,ember, provider, patient advocate/navigator)

• How have they been helpful?

• Did your initial reaction change over time?

• Did you learn more about your results since first getting them?

• If so, what did you learn?

• Who or where did you learn this from? (A friend, family member, healthcare

provider, genetic counselor, internet?)

- If internet, do you remember what sites you visited?

Now, I’d like to ask you about the impact your genetic test result had on your

family members.

• Do you think your results mean something to your family members’ risk of

developing breast cancer?

- If yes, which family members?

• Did you share your results with your family members? If so, who did you tell?

- Was telling others difficult? What was their reaction?

• What made the conversation difficult?

Now, I’d like to ask you about the impact your genetic test result has had on your

daily life.

• How has your genetic test result influenced your life, if at all?

• Has your result influenced your family relationships? Your plans for career or

family?

• What actions have you taken since receiving your genetic test result [state their

result]?

- For example, have you taken any new medications to reduce your breast cancer risk?
- Have you changed your screening habits for breast cancer?
- Have you changed you’re eating, exercise, smoking, drinking, or drug

use?

• What is your understanding of the known risk factors related to breast cancer?

- What do you understand about the genetics of breast cancer?
- What is your understanding of how breast cancer may run in families?
- Is it passed down in families?
- Do you have to have a family history of breast cancer to develop the disease?

• How do you feel about your genetic testing experience?

- Do you feel genetic testing met your needs? Why or why not?
- Do you have any recommendations on how the genetic testing process could be improved for others receiving a test result like the one you received?

*If intervention patient (know this in advance of the interview):*

Finally, I’d like to ask you a few questions about your experience with the use of RealRisks as part of this study.

• Did you have the chance to complete RealRisks as part of this study?

- If not, why not? (i.e., time constraints, uncomfortable with website features, technology issues?) If patient did complete RealRisks:

• Do you feel that RealRisks was helpful in increasing your education about

genetic testing for breast cancer?

- Why or why not?

• How did your completion of the modules about genetic testing in RealRisks

impact your decision to see a genetic counselor/get genetic testing?

• Looking back, is there anything you would change about the RealRisks website?

*Conclude the interview: thank the participant for their time and inform them*
